# Early viral clearance and antibody kinetics of COVID-19 among asymptomatic carriers

**DOI:** 10.1101/2020.04.28.20083139

**Authors:** Tongyang Xiao, Yanrong Wang, Jing Yuan, Haocheng Ye, Lanlan Wei, Xuejiao Liao, Haiyan Wang, Shen Qian, Zhaoqin Wang, Lei Liu, Zheng Zhang

**Affiliations:** Institute of Hepatology, National Clinical Research Center for Infectious Disease, Shenzhen Third People’s Hospital, Shenzhen 518112, Guangdong Province, China; Department of Pediatrics, National Clinical Research Center for Infectious Disease, Shenzhen Third People’s Hospital, Shenzhen 518112, Guangdong Province, China; Department of Infectious Diseases, National Clinical Research Center for Infectious Disease, Shenzhen Third People’s Hospital, Shenzhen 518112, Guangdong Province, China; The Second Affiliated Hospital, School of Medicine, Southern University of Science and Technology, Shenzhen 518112, Guangdong Province, China

**Keywords:** Asymptomatic, Viral clearance, Antibody, SARS-CoV-2, COVID-19

## Abstract

**Background:** Asymptomatic carriers contribute to the spread of Coronavirus Disease 2019 (COVID-19), but their clinical characteristics, viral kinetics, and antibody responses remain unclear.

**Methods:** A total of 56 COVID-19 patients without symptoms at admission and 19 age-matched symptomatic patients were enrolled. RNA of SARS-CoV-2 was tested using transcriptase quantitative PCR, and the total antibodies (Ab), IgG, IgA and IgM against the SARS-CoV-2 were tested using Chemiluminescence Microparticle Immuno Assay.

**Results:** Among 56 patients without symptoms at admission, 33 cases displayed symptoms and 23 remained asymptomatic throughout the follow-up period. 43.8% of the asymptomatic carriers were children and none of the asymptomatic cases had recognizable changes in C-reactive protein or interleukin-6, except one 64-year-old patient. The initial threshold cycle value of nasopharyngeal SARS-CoV-2 in asymptomatic carriers was similar to that in pre-symptomatic and symptomatic patients, but the communicable period of asymptomatic carriers (9.63 days) was shorter than pre-symptomatic patients (13.6 days). There was no obvious differences of the seropositive conversion rate of total Ab, IgG, and IgA among the three groups, though the rates of IgM varied largely. The average peak IgG and IgM COI of asymptomatic cases was 3.5 and 0.8, respectively, which is also lower than those in symptomatic patients with peaked IgG and IgM COI of 4.5 and 2.4 (p <0.05).

**Conclusion:** Young COVID-19 patients seem to be asymptomatic cases with early clearance of SARS-CoV-2 and low levels of IgM generation but high total Ab, IgG and IgA. Our findings provide empirical information for viral clearance and antibody kinetics of asymptomatic COVID-19 patients.

## Introduction

An outbreak of the 2019 novel coronavirus disease (COVID-19) has garnered international attention, rapidly spreading across the globe since it was first diagnosed in December of 2019. The World Health Organization (WHO) has confirmed more than 2.4 million COVID-19 cases worldwide, resulting in approximately 0.16 million deaths, as of April 21, 2020 (1). The infectiousness and transmission of the COVID-19 is particularly worrisome to public health officials, as a number of cases have spread asymptomatically. Asymptomatic transmission of SARS-COV-2 has been documented (2, 3), with the proportion of cases attributed to asymptomatic transmission varying widely, with rates ranging from 1.2% to 50% (4, 5). Asymptomatic infection refers to a person who has no clinical symptoms (such as fever, cough, or sore throat), yet test positive for the virus or serum antibody against severe acute respiratory syndrome coronavirus 2 (SARS-CoV-2) (6). SARS-CoV-2 has the possibility of viral shedding, thus transmission of the virus can occurs during the asymptomatic period (7–9). Patients can be infected and transmit the disease without showing symptoms, suggesting that perhaps further isolation and continuous nucleic acid testing may be warranted after a patient is discharged (3). As no vaccine has yet to be developed and treatment options are limited, identifying and containing the spread of these asymptomatic infections are key interventions that are necessary if governments and healthcare systems are to control the spread of COVID-19 and reduce disease related morality.

Because asymptomatic patients are difficult to track and find, they may contribute to the spread of infection and even lead to secondary outbreaks. Therefore, the screening and diagnosing asymptomatic carriers, particularly in the early phase of infection, is beneficial for the prevention and treatment of COVID-19. Asymptomatic infections may be flare up due to weakened immune responses and subclinical manifestations, or because the virus is waiting for opportunities to reproduce and invade. Thus, the aim of this study was to understand its mechanism, viral clearance, and antibody kinetics of asymptomatic carriers in order to better inform national control policies and prevent further infection.

## Methods

### Study design and participants

This was a retrospective analysis of patients with confirmed COVID-19 from the Third People’s Hospital of Shenzhen between January 23, 2020 and April 1, 2020 were enrolled in this study. Inclusion criteria were patients who tested positive for the SARS-CoV-2 virus or had serum antibodies to the virus. Cases were divided into asymptomatic, pre-symptomatic, or symptomatic cases based on the patients’ symptoms and disease-related outcomes. The asymptomatic and pre-symptomatic patients enrolled in this study were identified through active surveillance and contact tracing. Asymptomatic carriers showed no self-perceived or clinically recognizable symptoms from admission until 14 days post-discharge. Pre-symptomatic patients had no symptoms at admission, but gradually showed symptoms such as fever, cough, chest discomfort, diarrhea, headache, and myalgia during hospitalization. Symptomatic patients showed symptoms such as fever, cough, chest discomfort, diarrhea, headache, and myalgia at admission. The discharge criteria for recovered patients was: 1) normal temperature for more than 3 days; 2) respiratory symptoms significantly improved, classified as significant absorption of pulmonary lesions on chest computerize tomography (CT) scans; 3) two consecutive negative ribonucleic acid (RNA) tests in 24 hours. Abnormal chest CT was the presence of unilateral lobe lesions, multiple lobes in both lungs, or all lobes in both lungs. Finally, 23 asymptomatic, 33 presymptomatic and 19 age-matched symptomatic patients was selected. This study was approved by the Ethics Committee of Shenzhen Third People’s Hospital (2020-169), which waived the requirement for written patient consent for this retrospective analysis. All patients gave their oral consent to participate in this study.

### Data collection

The medical records of 449 COVID-19 patients were reviewed. The epidemiological, demographic, clinical, and laboratory data of these patients were retrospectively collected. According to the Guidelines for the Diagnosis and Treatment for Novel Coronavirus Pneumonia (the Sixth Edition) published by National Health Commission of the People’s Republic of China (6), all diagnosed cases of COVID-19 were confirmed using respiratory reverse transcription polymerase chain reaction (RT-PCR) tests or antibody tests for SARS-CoV-2.

Ribonucleic acid (RNA) was extracted from 200 μL of respiratory or anal swab specimens using the Huayin-Bio Viral RNA Mini-Kit (Huayin, Shenzhen, China). Samples for SARS-CoV-2 virus were tested using the RT-PCR Kit (GeneoDX Co., Ltd., Shanghai, China) on an ABI 7500 thermo cycler. Only the runs with valid internal reference were included. Each RT-PCR assay provided a threshold cycle (Ct) value, which is the number of cycles required for the fluorescent signal to cross the threshold for a positive test, with a higher Ct value correlated to a lower viral load. The specimens were considered positive if the Ct value was < 40.0, and negative if the viral load was undetectable. Specimen testing was repeated if the cycle-threshold value was higher than 37. The specimen was then considered positive if the repeat results were the same as the initial result and between 37 and 40. If the repeat Ct was undetectable, the specimen was considered negative. All procedures involving clinical specimens and SARS-CoV-2 were performed in a Biosafety Level 3 Laboratory.

The SARS-CoV-2 specific total antibody (Ab), IgG, IgA, and IgM in plasma was tested using a Chemiluminescence Microparticle Immuno Assay (CMIA). Briefly, antigens containing the receptor-binding domain (RBD) were used as the immobilized and HRP-conjugated antigen to detect total antibodies by double-antigens sandwich enzyme-linked immunosorbent assay (Ab-ELISA). IgM was tested by the IgM μ-chain capture method (IgM-ELISA), using the same RBD antigen as the Ab-ELISA. IgA and IgG were tested by the indirect ELISA using RBD antigen. The testing kits were supplied by the Beijing Wantai Biological Pharmacy Enterprise Co., Ltd., China. Fluorescence intensity was used to measure antibody concentration. The relative fluorescence of sample to control (COI) was used to estimate the result. When COI was more than one, the result was judged to be positive.

### Statistical Analysis

The log2 transformation was done for COI. Variables were described using median, interquartile range (IQR), mean ± standard deviation (SD), or percentages. Chi-square or Fisher’s exact tests were utilized to compare the proportions of the categorical variables. One-way ANOVA or Wilcoxon rank-sum tests were used for the continuous variables. The statistical tests were two-sided, and significant differences were considered at *P* < 0.05. All statistical analysis was performed using GraphPad Prism 8.3.0 (GraphPad Software, La Jolla, California, USA, www.graphpad.com).

## Results

### Demographic characteristics of Asymptomatic SARS-CoV-2 infected patients

Figure 1 summarizes the study design. From January 11 to April 1, 2020, a total of 449 patients (417 native patients and 32 imported cases) were admitted to the Third People’s Hospital of Shenzhen. 77 cases (17.15%) showed no symptoms at admission, but 21 cases were excluded due to severe conditions (n = 2), inpatients (n = 5), or undetectable RNA and IgM (n = 14). 56 patients whom were discharged and without symptoms at admission were enrolled in the final analysis. Of the 372 cases with symptoms at admission, 19 were aged-matched moderate patients whom were selected as controls. Finally, 23 asymptomatic, 33 pre-symptomatic, and 19 age-matched symptomatic patients was selected.

**Figure 1.**
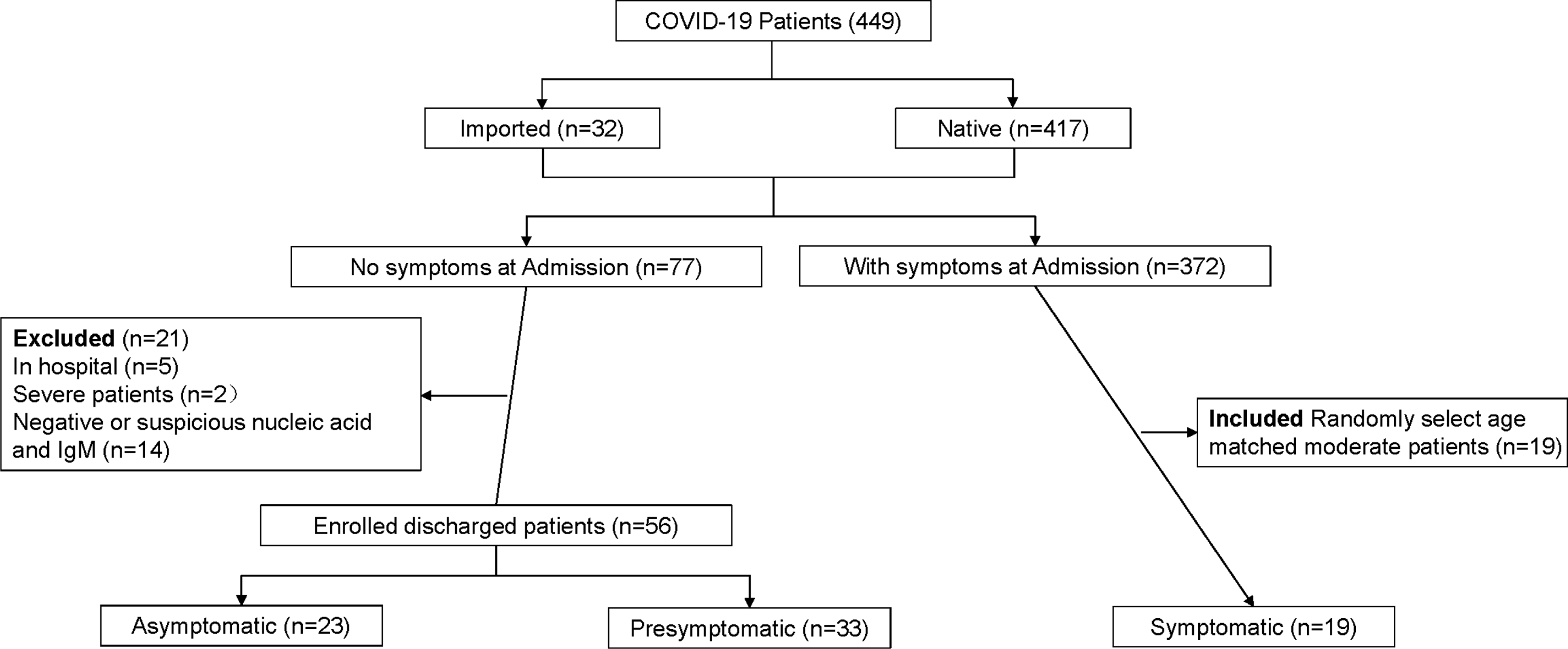
Participant profile. The figure depicts the study design and general inclusion criteria of the study.

### Clinical characteristics of asymptomatic patients

As seen in Table 1, asymptomatic carriers were younger compared to pre-symptomatic and symptomatic patients. 43.5% of asymptomatic carriers were children, of whom two are infants, 78.3% were female, and two (8.7%) had comorbidities, such as hypertension and asthma. 34.8% of asymptomatic carriers and 30.3% of pre-symptomatic patients tested positive for SARS-CoV-2 RNA after discharge. 56.5% of asymptomatic carriers showed abnormal manifestations, while more than 80% of pre-symptomatic patients had abnormal chest CT images, such as ground-glass opacity and bilateral patchy shadowing (p < 0.05). Compared with symptomatic patients, the number of days from symptom onset to hospitalization was shorter (p < 0.05) for asymptomatic and pre-symptomatic carriers. However, the length of hospital stay for asymptomatic and pre-symptomatic patients was slightly longer compared to symptomatic patients, but was not found to be signficant (p = 0.07). All participants received antiviral drugs such as Ritonavir, Chloroquine, Ribavirin, Arbidol, and interferon.

**Table 1.**
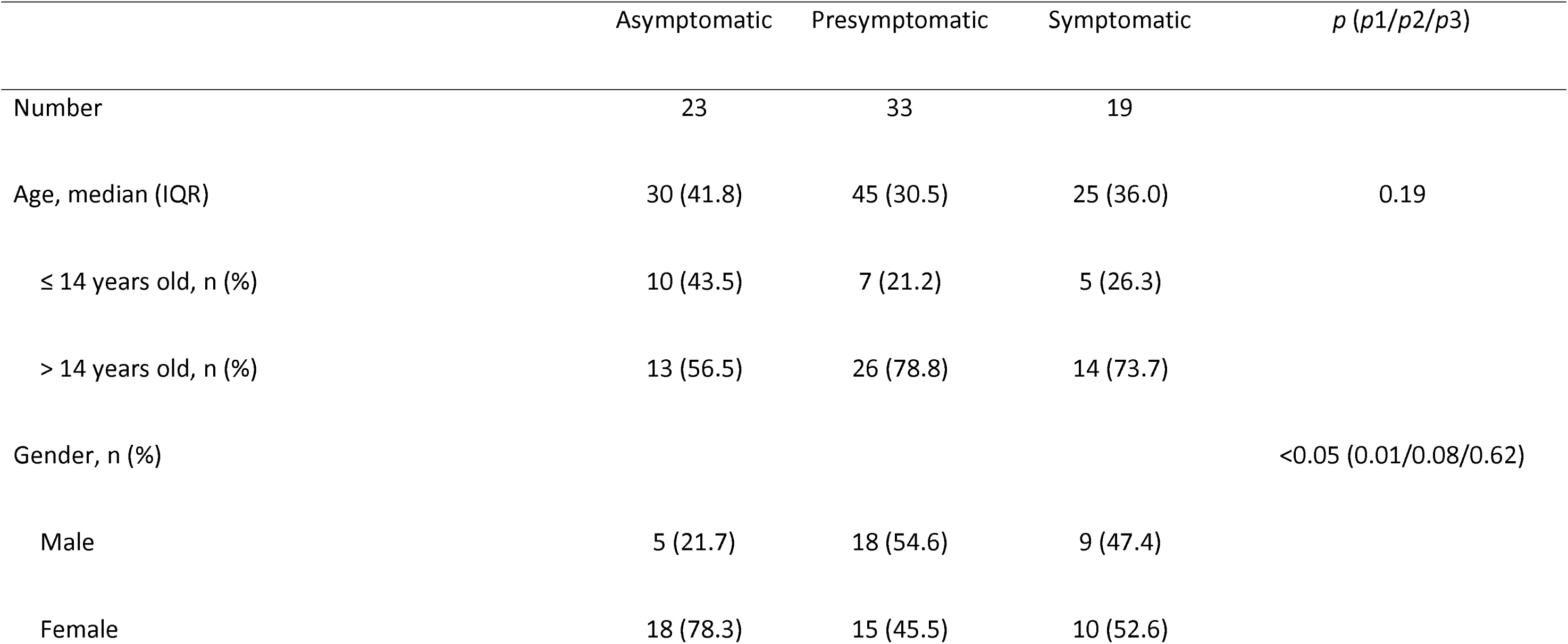
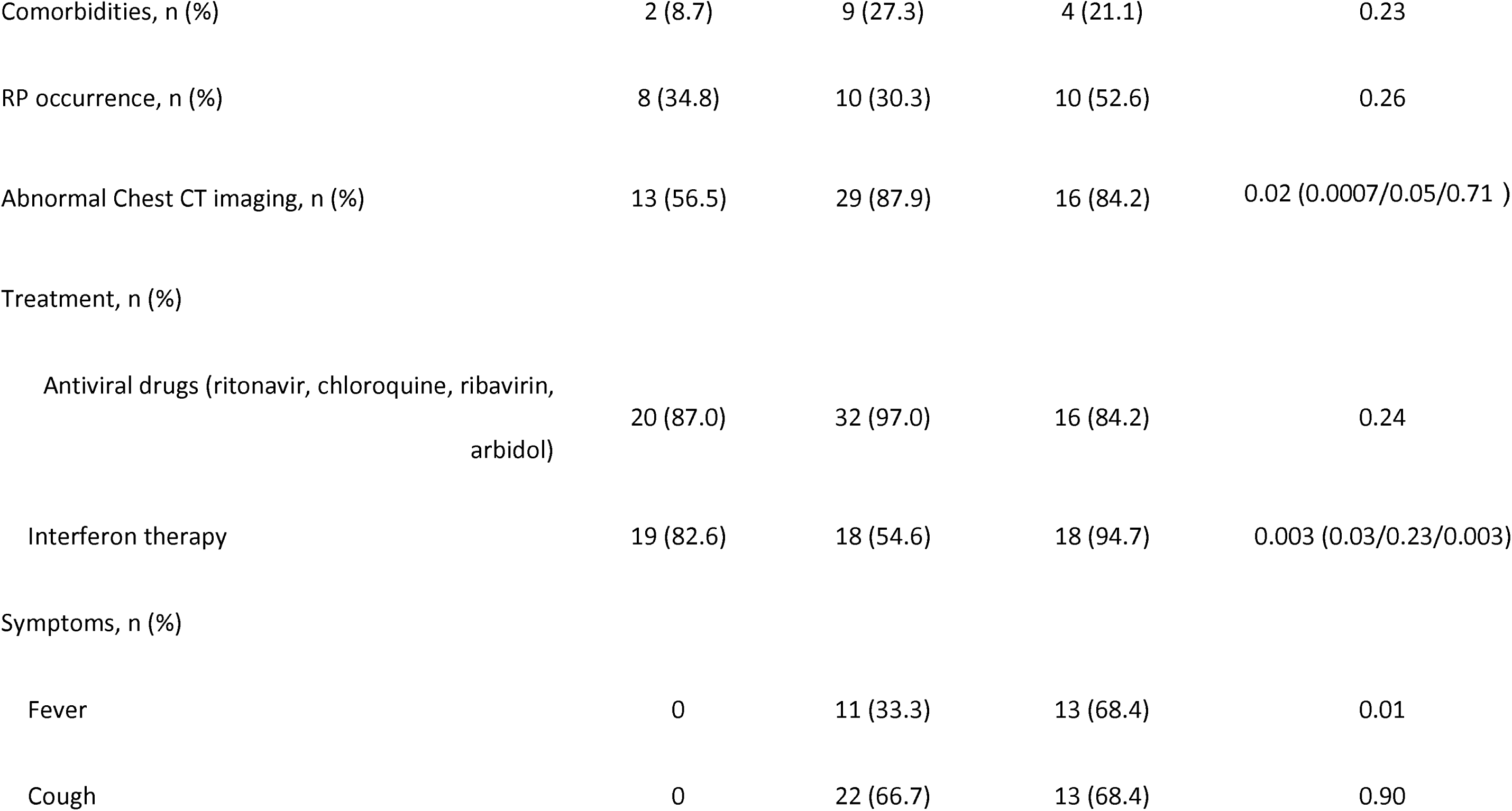
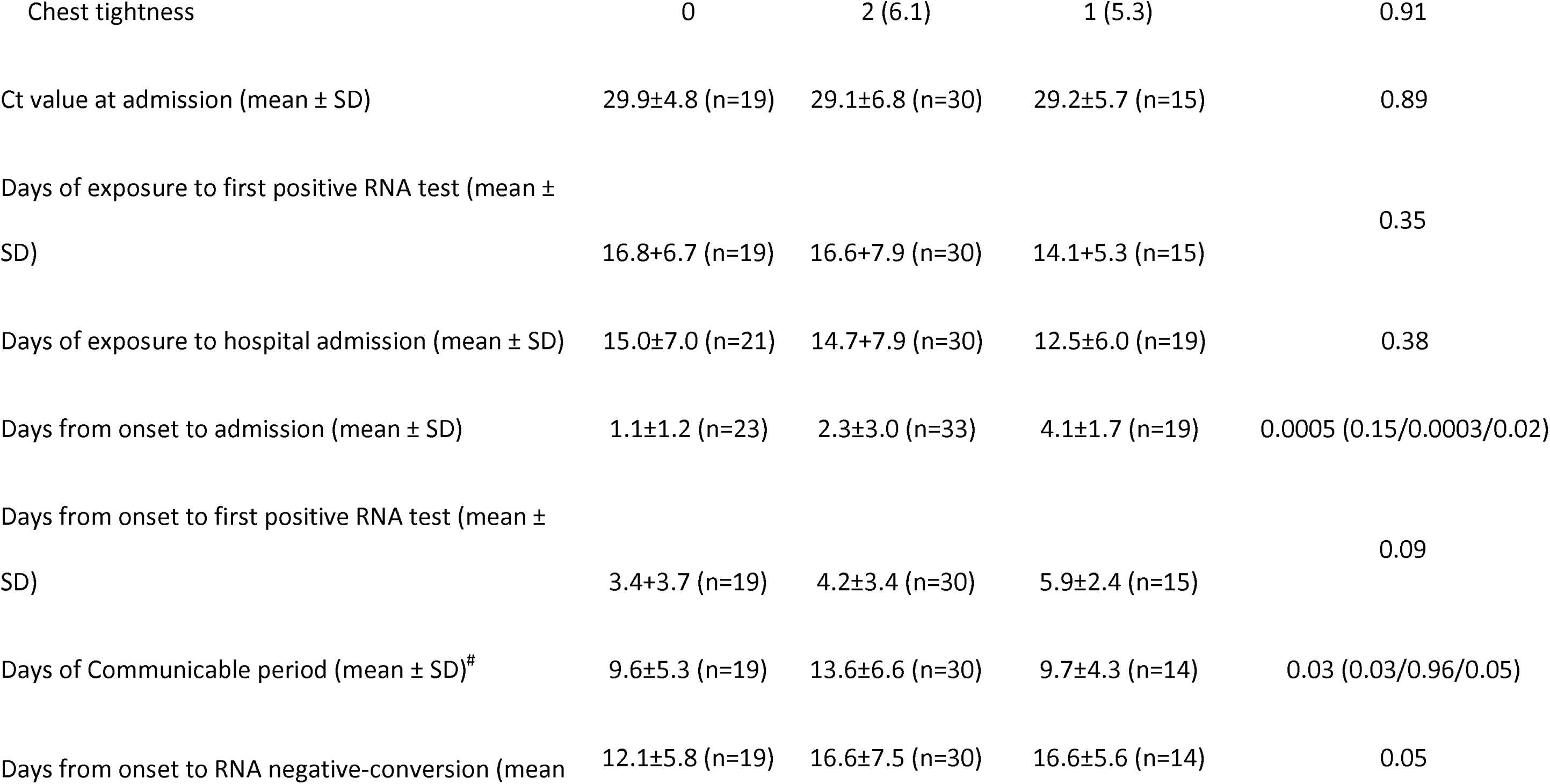
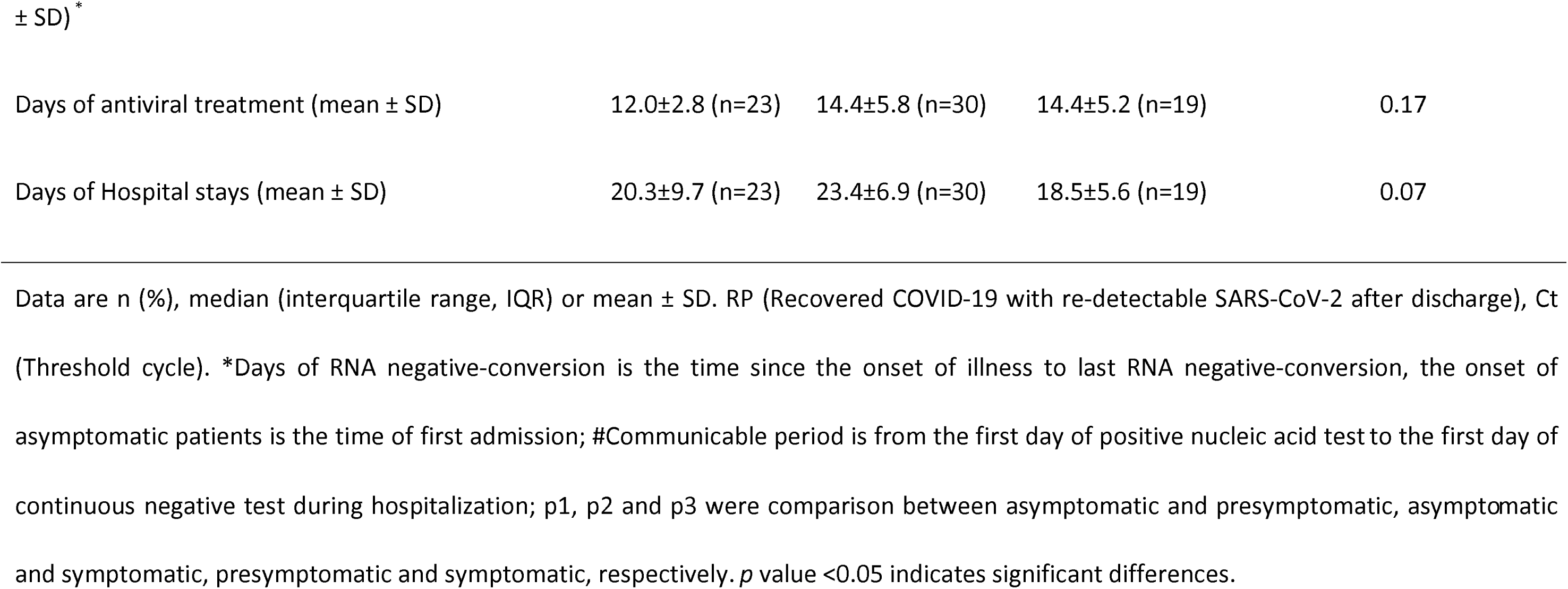
Clinical information of the enrolled patients

|  | Asymptomatic | Presymptomatic | Symptomatic | *p* (*p*1/*p*2/*p*3) |
| --- | --- | --- | --- | --- |
| Number | 23 | 33 | 19 |  |
| Age, median (IQR) | 30 (41.8) | 45 (30.5) | 25 (36.0) | 0.19 |
| ≤ 14 years old, n (%) | 10 (43.5) | 7 (21.2) | 5 (26.3) |  |
| > 14 years old, n (%) | 13 (56.5) | 26 (78.8) | 14 (73.7) |  |
| Gender, n (%) |  |  |  | <0.05 (0.01/0.08/0.62) |
| Male | 5 (21.7) | 18 (54.6) | 9 (47.4) |  |
| Female | 18 (78.3) | 15 (45.5) | 10 (52.6) |  |
| Comorbidities, n (%) | 2 (8.7) | 9 (27.3) | 4 (21.1) | 0.23 |
| RP occurrence, n (%) | 8 (34.8) | 10 (30.3) | 10 (52.6) | 0.26 |
| Abnormal Chest CT imaging, n (%) | 13 (56.5) | 29 (87.9) | 16 (84.2) | 0.02 (0.0007/0.05/0.71 ) |
| Treatment, n (%) |  |  |  |  |
| Antiviral drugs (ritonavir, chloroquine, ribavirin, arbidol) | 20 (87.0) | 32 (97.0) | 16 (84.2) | 0.24 |
| Interferon therapy | 19 (82.6) | 18 (54.6) | 18 (94.7) | 0.003 (0.03/0.23/0.003) |
| Symptoms, n (%) |  |  |  |  |
| Fever | 0 | 11 (33.3) | 13 (68.4) | 0.01 |
| Cough | 0 | 22 (66.7) | 13 (68.4) | 0.90 |
| Chest tightness | 0 | 2 (6.1) | 1 (5.3) | 0.91 |
| Ct value at admission (mean ± SD) | 29.9±4.8 (n=19) | 29.1±6.8 (n=30) | 29.2±5.7 (n=15) | 0.89 |
| Days of exposure to first positive RNA test (mean ± SD) | 16.8±6.7 (n=19) | 16.6±7.9 (n=30) | 14.1±5.3 (n=15) | 0.35 |
| Days of exposure to hospital admission (mean ± SD) | 15.0±7.0 (n=21) | 14.7±7.9 (n=30) | 12.5±6.0 (n=19) | 0.38 |
| Days from onset to admission (mean ± SD) | 1.1±1.2 (n=23) | 2.3±3.0 (n=33) | 4.1±1.7 (n=19) | 0.0005 (0.15/0.0003/0.02) |
| Days from onset to first positive RNA test (mean ± SD) | 3.4±3.7 (n=19) | 4.2±3.4 (n=30) | 5.9±2.4 (n=15) | 0.09 |
| Days of Communicable period (mean ± SD)<sup>#</sup> | 9.6±5.3 (n=19) | 13.6±6.6 (n=30) | 9.7±4.3 (n=14) | 0.03 (0.03/0.96/0.05) |
| Days from onset to RNA negative-conversion (mean | 12.1±5.8 (n=19) | 16.6±7.5 (n=30) | 16.6±5.6 (n=14) | 0.05 |
| ± SD) \* |  |  |  |  |
| Days of antiviral treatment (mean ± SD) | 12.0±2.8 (n=23) | 14.4±5.8 (n=30) | 14.4±5.2 (n=19) | 0.17 |
| Days of Hospital stays (mean ± SD) | 20.3±9.7 (n=23) | 23.4±6.9 (n=30) | 18.5±5.6 (n=19) | 0.07 |

Data are n (%), median (interquartile range, IQR) or mean ± SD. RP (Recovered COVID-19 with re-detectable SARS-CoV-2 after discharge), Ct (Threshold cycle). \*Days of RNA negative-conversion is the time since the onset of illness to last RNA negative-conversion, the onset of asymptomatic patients is the time of first admission; #Communicable period is from the first day of positive nucleic acid test to the first day of continuous negative test during hospitalization; p1, p2 and p3 were comparison between asymptomatic and presymptomatic, asymptomatic and symptomatic, presymptomatic and symptomatic, respectively. *p* value <0.05 indicates significant differences.

### Laboratory characteristics of asymptomatic patients

Only a few asymptomatic cases had recognizable changes in laboratory tests (Table 2). C-reactive protein (CRP) and interleukin (IL)-6 are two inflammatory markers. At admission, all asymptomatic patients showed normal CRP and IL-6 levels which did not progress, except in one 64-year-old patient whose CRP and IL-6 were slightly increased until discharge. However, elevated CRP occurred in 8 pre-symptomatic and 6 symptomatic participants (p < 0.05) and increased IL-6 occurred in 9 pre-symptomatic and 8 symptomatic participants, despite both obtaining normal CRP and IL-6 after treatment (p < 0.05). Lactate dehydrogenase (LDH) was used as an indicator of disease severity. During the course of disease, LDH was increased in all of the groups and no significant difference were present in other laboratory tests, such myohemoglobin (MYO), oxyhemoglobin saturation (O_2_AT) between the three groups. In addition, the average white blood cell (WBC), lymphocyte, CD4+ T cell, and CD8+ T cell counts were slightly changed within normal ranges during the hospitalization period, but no significant difference was observed among the three groups (Figure 2).

**Table 2.**
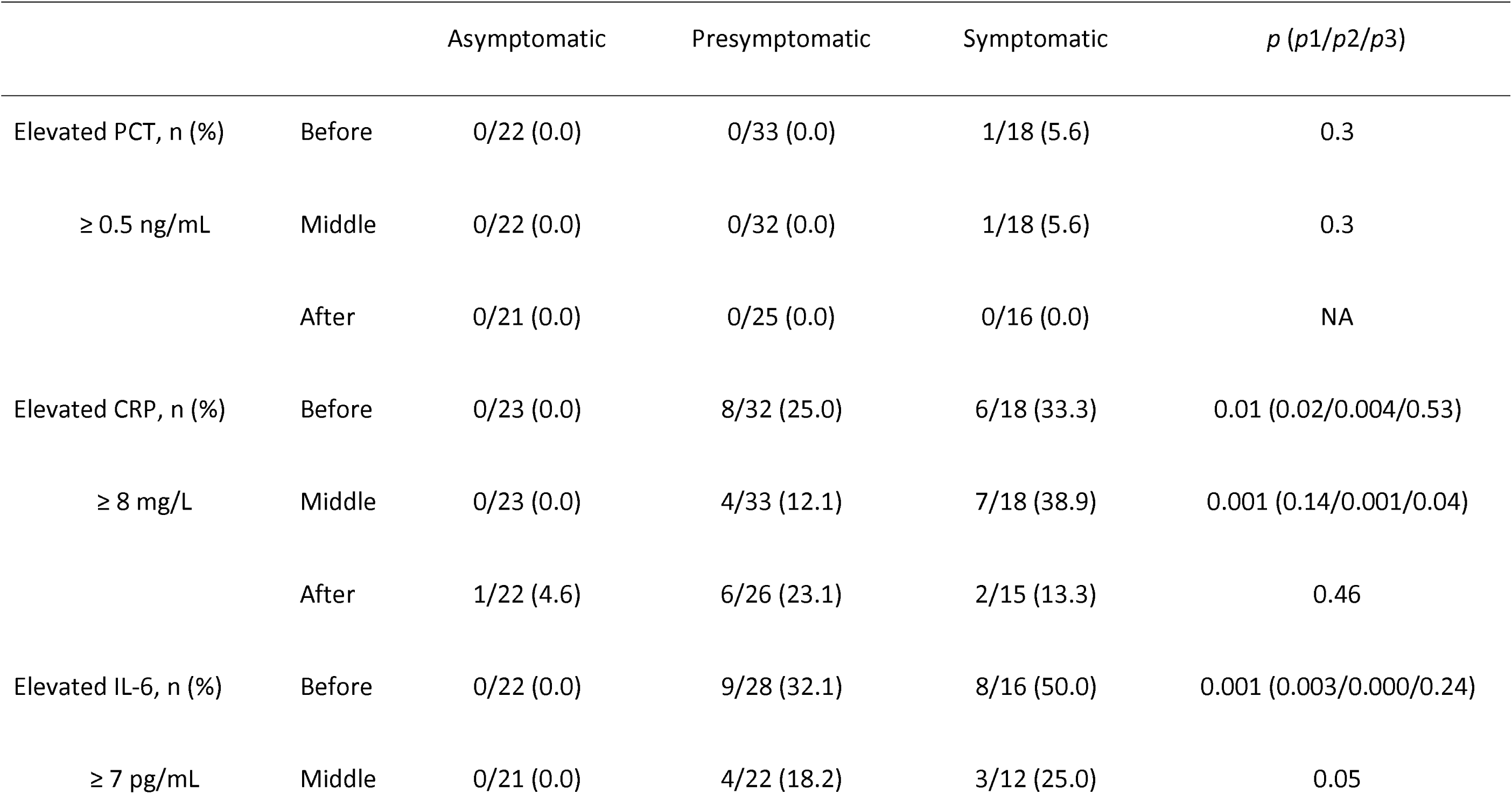
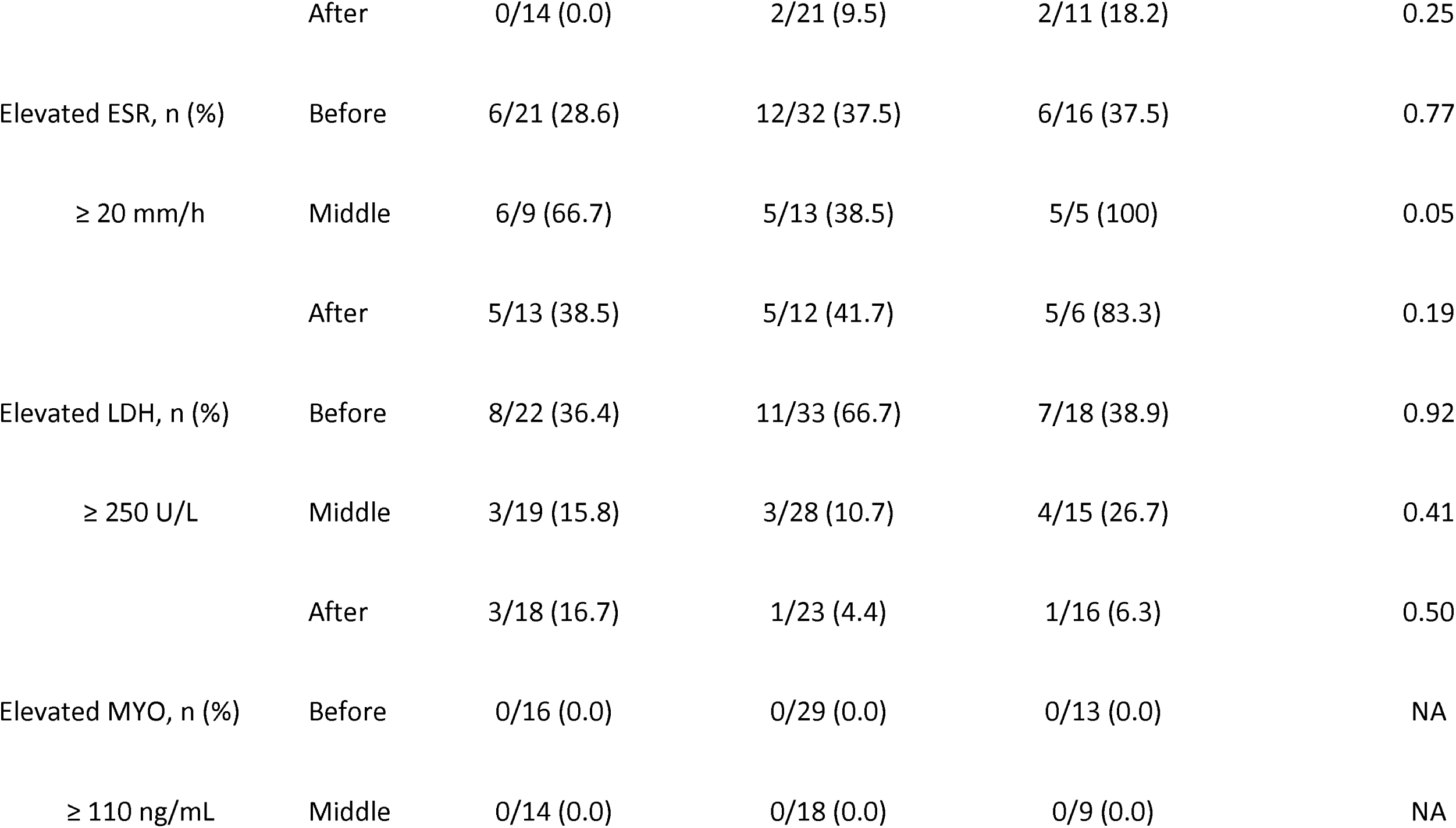
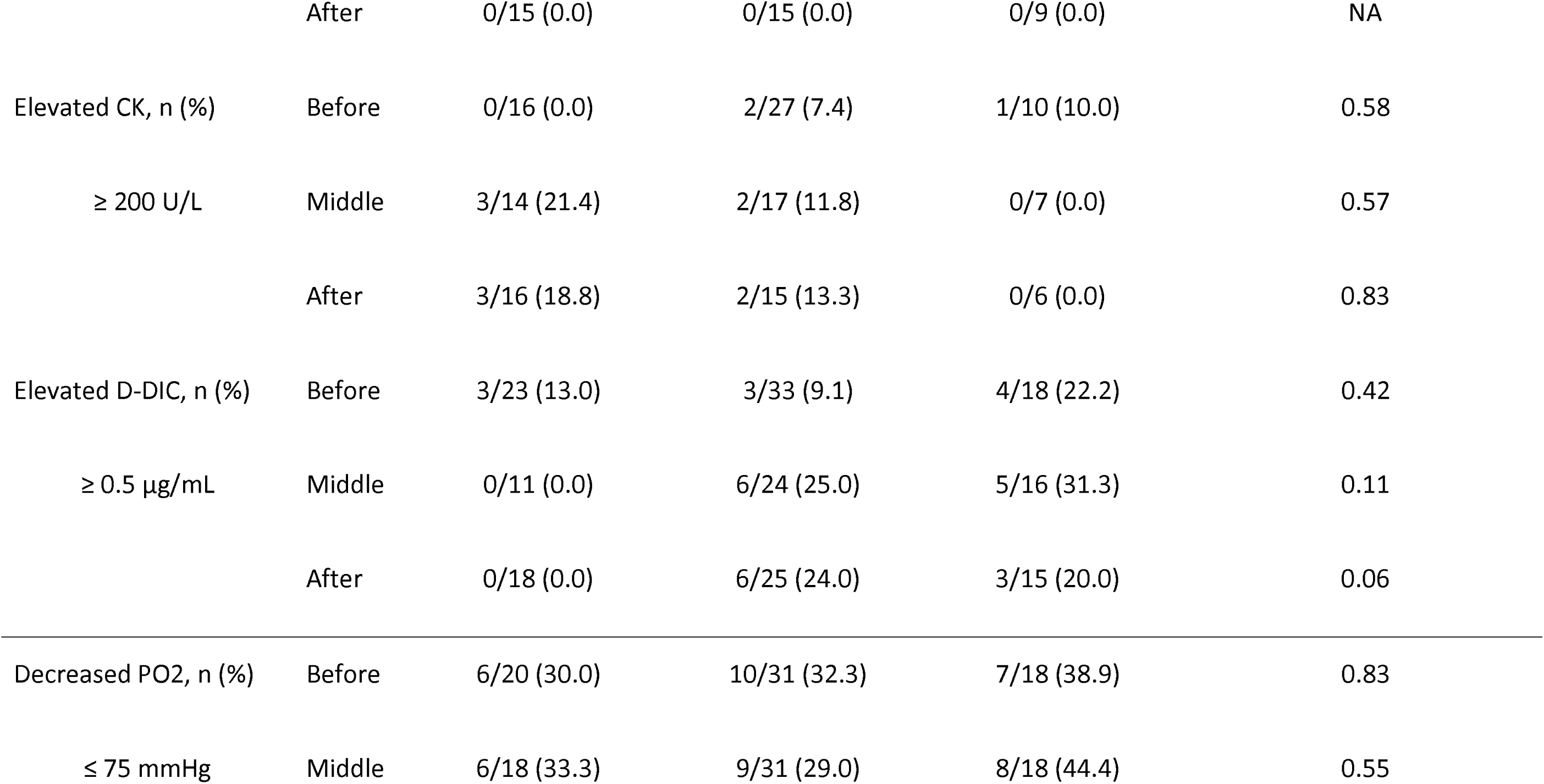
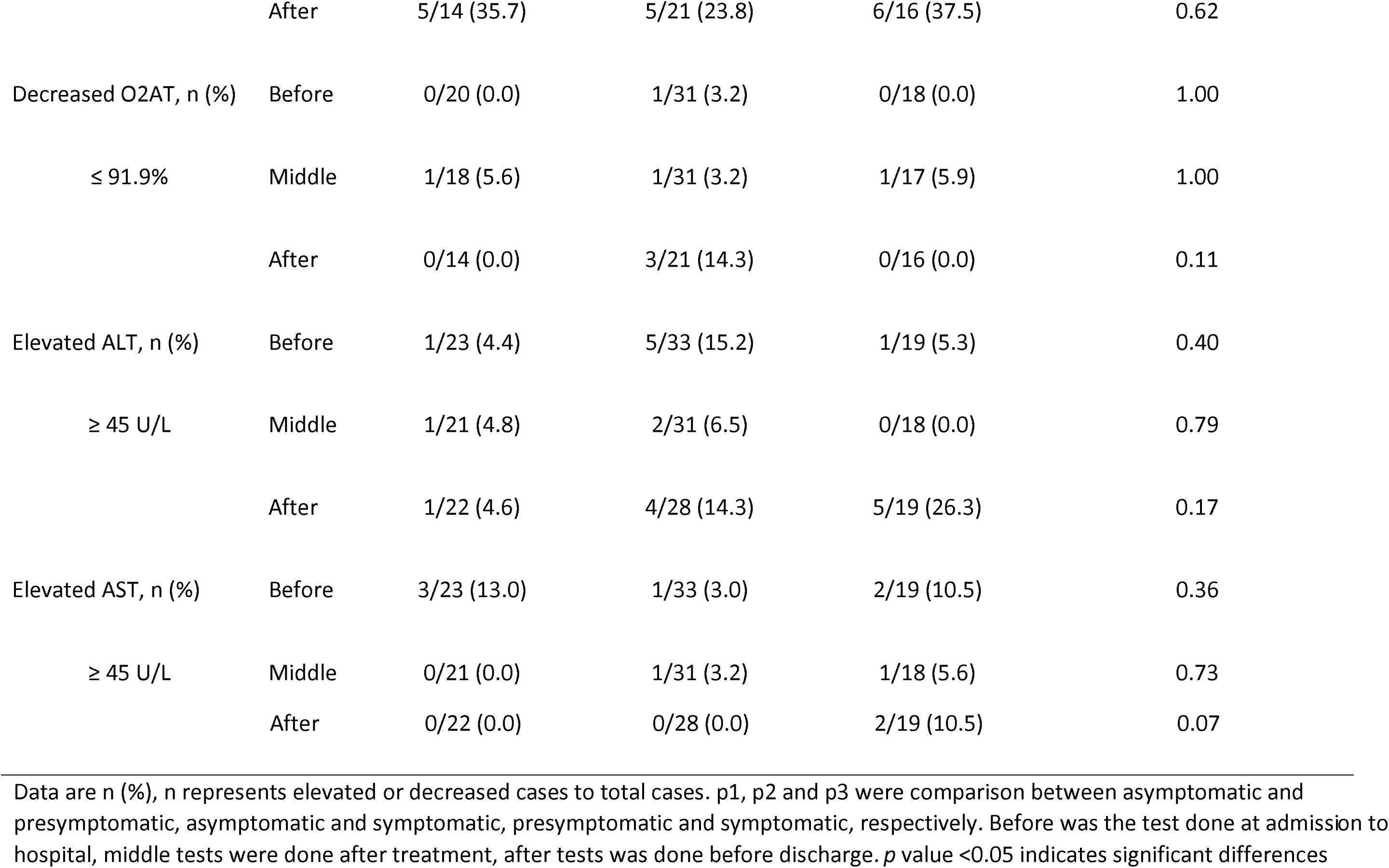
Laboratory characteristics of asymptomatic patients

**Figure 2.**
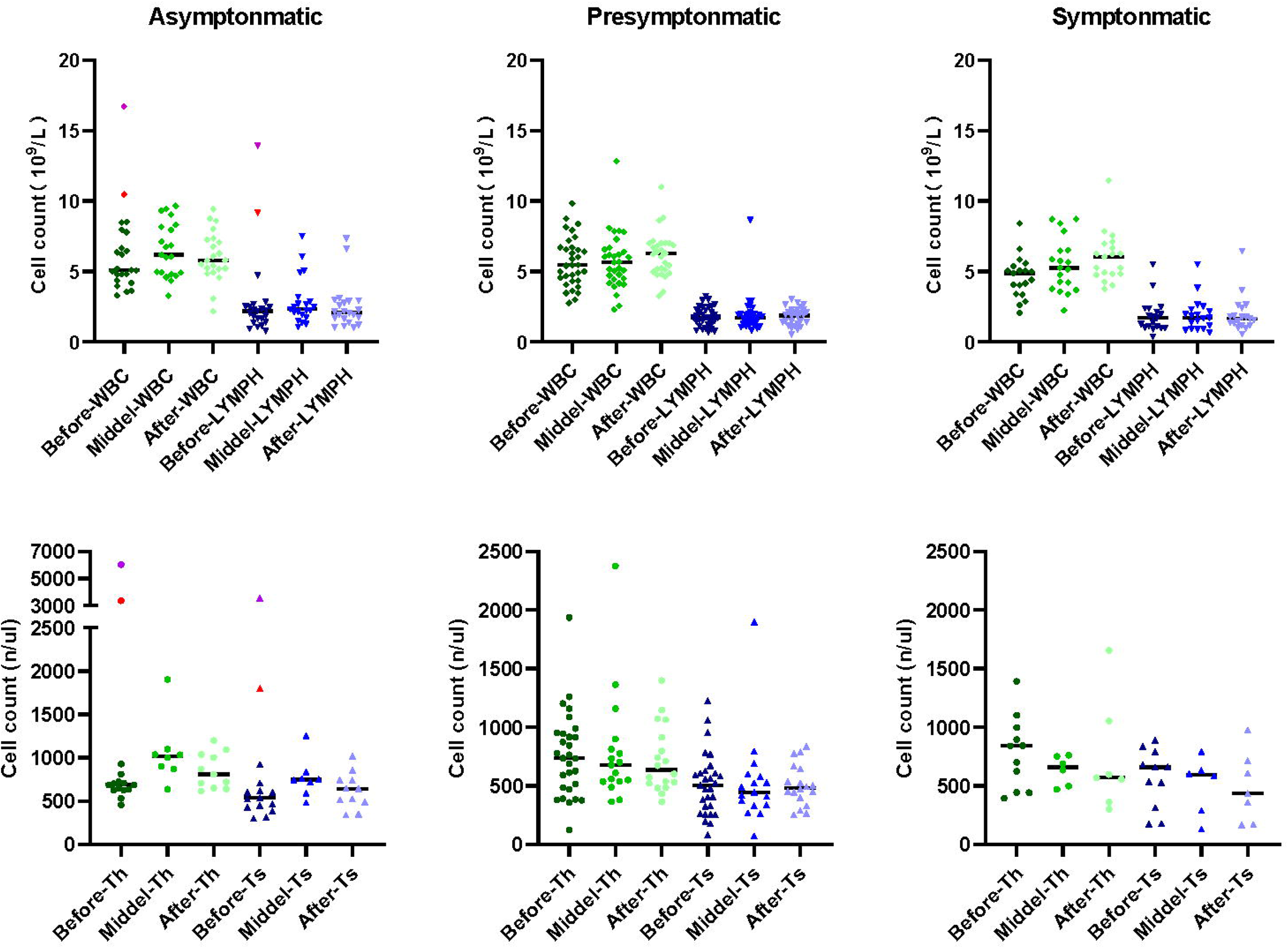
Dynamics of WBC, lymphocyte, CD4+ T, and CD8+ T cell counts. The total number of white blood cells (WBC), lymphocytes (LYMPH), CD4+ T, and CD8+ T cell tests at admission (Before) to hospital, after treatment (Middle) and before discharge (After) was shown. The red and purple dots represent two infants enrolled in this study.

### Rapid viral clearance in asymptomatic cases

There was no obvious difference in the calculated initial Ct value of nasopharyngeal samples among the three groups (Table 1). The average communicable duration of asymptomatic cases was 9.63 days, which was significantly shorter than 13.6 days in pre-symptomatic patients (p < 0.05). Interestingly, the symptomatic patients also had a short communicable period (9.71 days). Similarly, the days of RNA negative-conversion of asymptomatic patients was slightly short than other two groups. The average conversion days of asymptomatic, presymptomatic and symptomatic was 12.11, 16.63 and 16.64, respectively.

The kinetics of SARS-CoV-2 RNA in nasopharyngeal and anal samples are shown in Figures 3A and 3B, respectively. The model-based initial viral load in nasopharyngeal samples of asymptomatic cases was lower, with more than 30 Ct values, as compared to pre-symptomatic and symptomatic cases, which had less than 30 Ct values. The RNA negative-conversion of asymptomatic cases occurred within 15–20 days after the onset and occurred after 20 days from the onset of admission in pre-symptomatic and symptomatic patients. On the contrary, the initial viral load in anal samples of asymptomatic cases was slightly higher than that in pre-symptomatic and symptomatic patients, and the RNA negative-conversion time was longer than in pre-symptomatic patients. In addition, the re-appearance of SARS-CoV-2 RNA could be observed in nasopharyngeal and anal samples 54 and 42 days, respectively, after the onset of admission for some asymptomatic carriers, 81 and 40 days for pre-symptomatic patients, and 62 and 42 for symptomatic patients, respectively.

**Figure 3.**
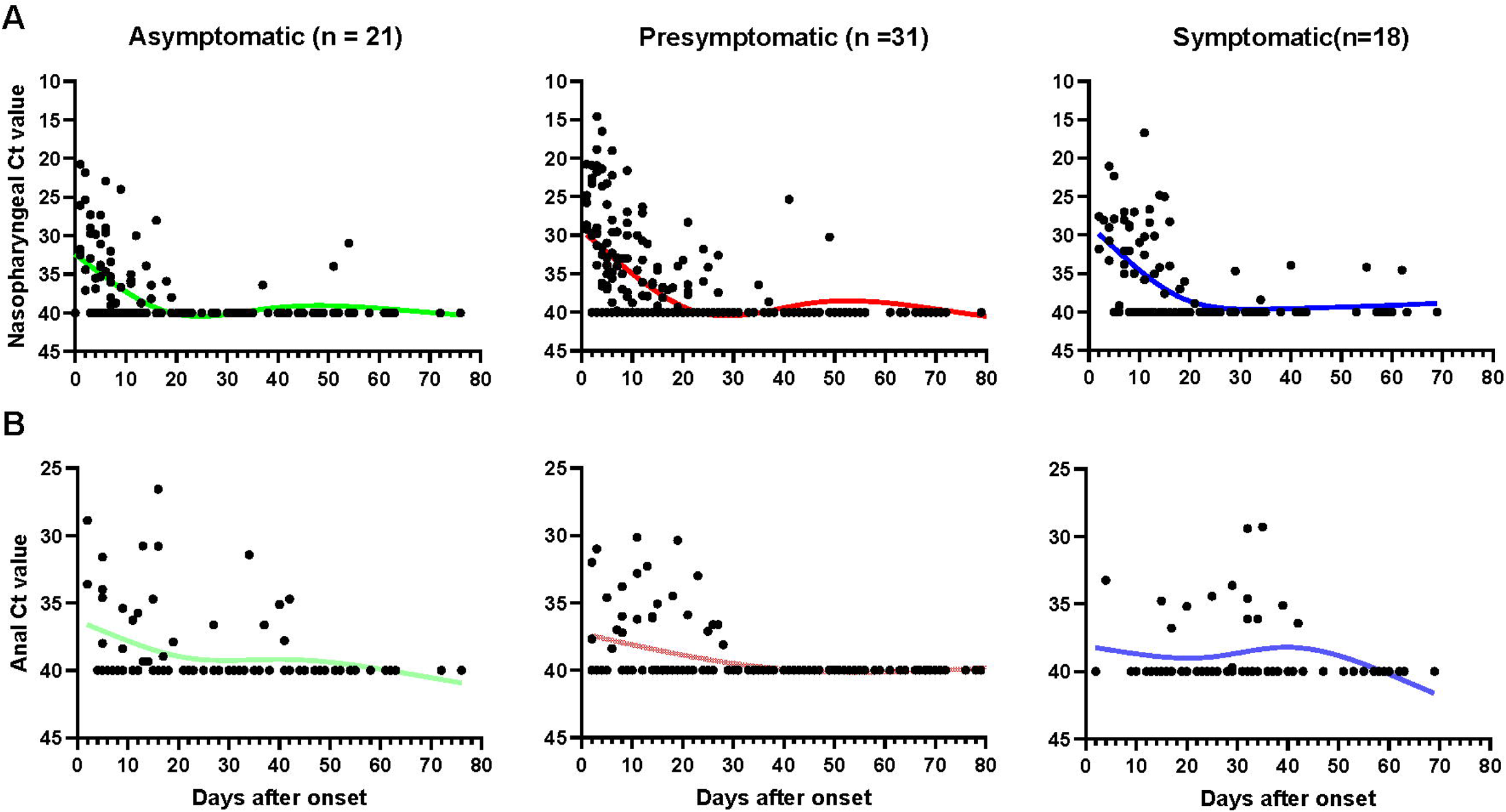
The kinetics of SARS-CoV-2 RNA in nasopharyngeal and anal samples. Threshold cycle (Ct) of nasopharyngeal (A) and anal (B) samples from patients with positive SARS-CoV-2 RNA test. Patients with totally negative results were not enrolled. Each point represents a sample, curves represent best fit line. Negative results are denoted with a Ct of 40.

### Sero-conversion of antibodies against SARS-CoV-2

A total of 324 plasma samples were detected for total Ab, IgM, IgG, and IgA against SARS-CoV-2, including samples from 77 asymptomatic carriers, 142 pre-symptomatic patients, and 105 symptomatic patients. The total sero-positive conversion rate for Ab, IgG, and IgA of asymptomatic patients was 90.9% (20/22), 95.5% (21/22) and 90.9% (20/22), respectively (Table S1); pre-symptomatic patients with 93.8% (30/32), 93.8% (30/32) and 90.6% (29/32); and all symptomatic patients experienced sero-positive conversion of total Ab, IgG and IgA. As shown in Figure 4, there was no obvious difference of total Ab, IgG, and IgA among the three groups (p > 0.05). Nearly half of cases had total Ab, IgG, and IgA within 1-7 days since the onset and more cases had antibodies within 8–14 days then almost all of cases had these antibodies within 15–30 days and were maintained with time (Figure 4A, 4B, 4D). Notably, none of the asymptomatic carriers loss IgA, while 6 (21.4%) of pre-symptomatic and 3 (15.8%) symptomatic patients lost IgA during follow-up.

**Figure 4.**
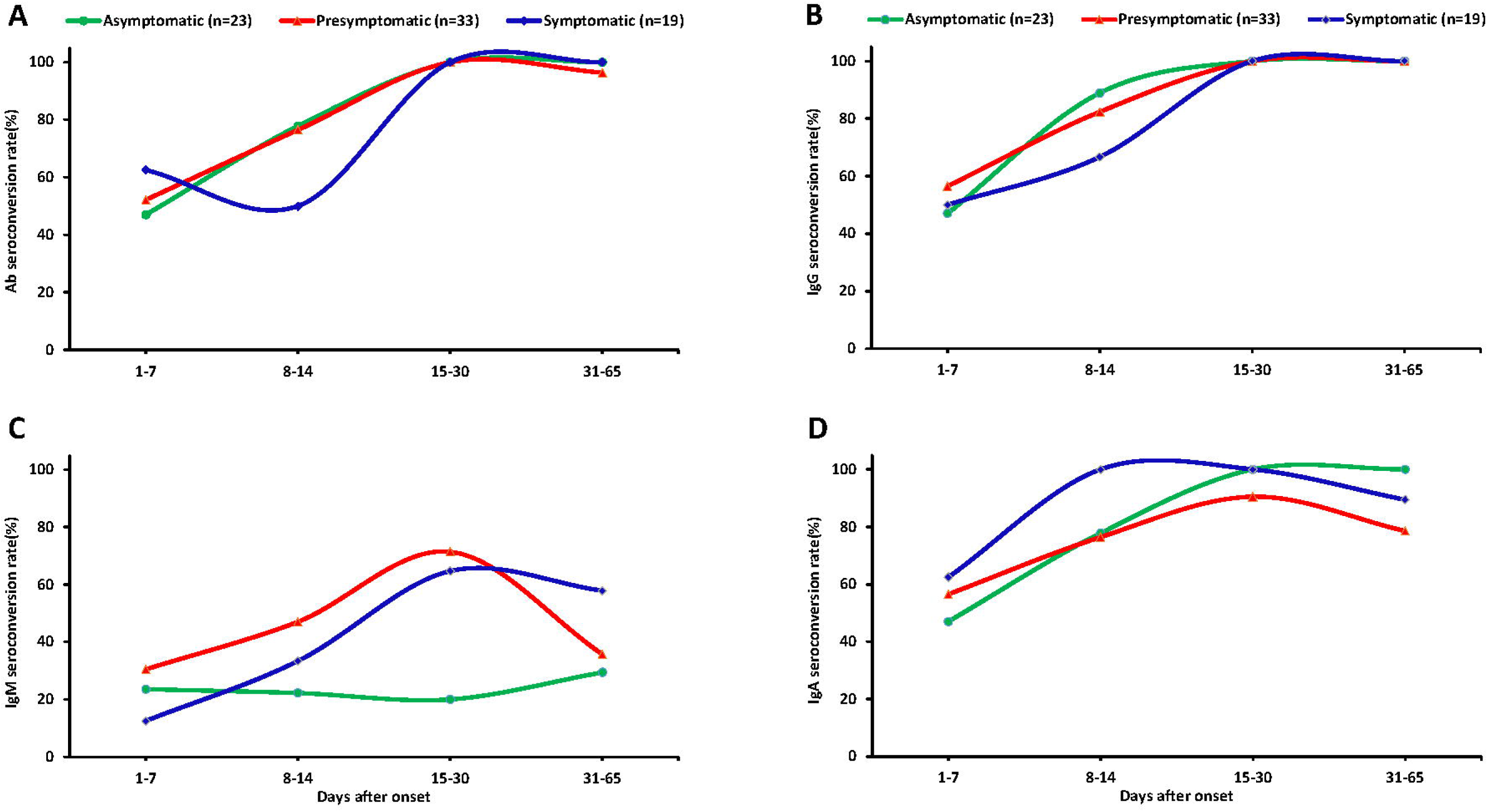
Seroconversion rate of antibodies in plasma of different patients. Dynamic seropositive rate of total Ab (A), IgG (B), IgM (C) and IgA (D) of different patients at different stage. Patients were divided into four age groups: 1–7 years, 8–14 years, 15–30 years.

The total seropositive conversion rate of IgM reached 45.5% (10/22), 62.5% (20/32), and 63.2% (12/19) for asymptomatic, pre-symptomatic, and symptomatic patients (Table S1), respectively. Importantly, IgM varied widely among the three groups. In pre-symptomatic and symptomatic patients, 30.4% and 12.5% of cases had IgM within the first week after onset of illness and the cases with IgM were increased until their peak 15 to 30 days after onset of illness (71.4% and 64.7%) and were then gradually decreased. The seropositive conversion rate of IgM decreased to 55% and 41.7% for pre-symptomatic and symptomatic patients, respectively, at one to two months after admission. By contrast, only 20% of asymptomatic cases produced IgM during the first week of admission, and most of cases failed to produced IgM at subsequent follow-ups (Figure 4C).

### Dynamics of antibody response with time

The IgG peaked as early as 20 days after the onset of admission for asymptomatic cases, but the pre-symptomatic and symptomatic patients peaked later (Figure 5A-5D). The average number of days until peak IgG COI of asymptomatic cases was 3.5 days, which was lower than symptomatic patients, which peaked at COI of 4.5 (p < 0.05, Table S2). The IgG was found to last in all three groups for two months or longer. Similar dynamics occurred for total Ab responses in the three groups (Figure 5A). Most of the asymptomatic cases had undetectable IgM, with concentrations varying slightly over time. Pre-symptomatic and symptomatic patients had higher IgM levels and some of them had persistent IgM levels for more than 70 days (Figure 5C). Compared with symptomatic patients, the peak IgM was significantly lower, with COI of 0.84, in asymptomatic cases (p < 0.05, Table S2). No obvious differences were observed in the dynamics of IgA among the three groups (Figure 5D), though the IgA peaked sooner in asymptomatic and pre-symptomatic patients, compared to symptomatic participants.

**Figure 5.**
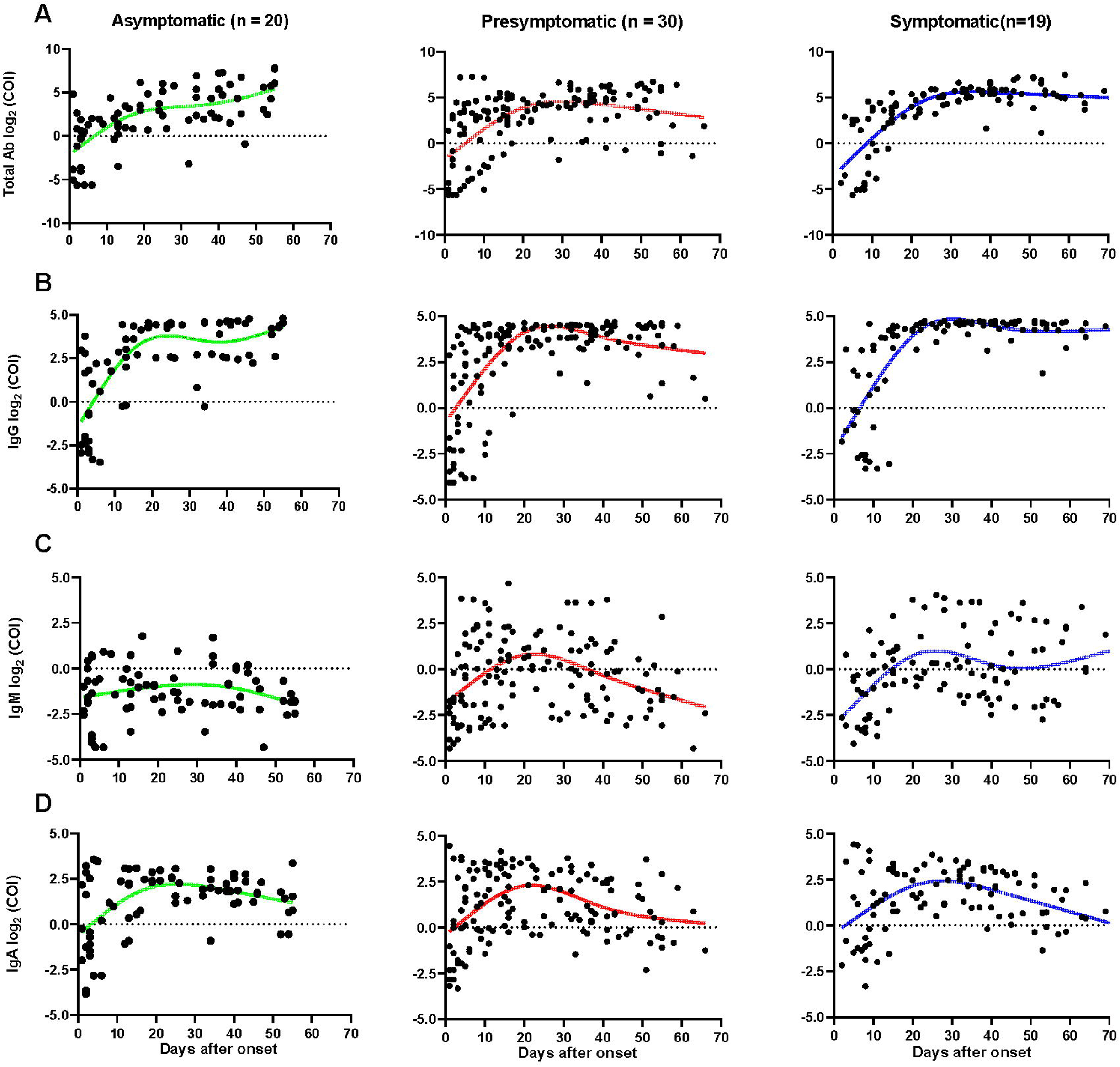
Dynamics of SARS-CoV-2 specific antibodies. The levels of total Ab (A), IgG (B), IgM (C), and IgA (D) of different patients after onset. The relative antibody level was estimated using log_2_ (COI). Each dot represents a sample, curves represent best fit line. Patients with totally negative antibodies or only one sample was excluded. Negative results are shown below the dotted horizontal lines.

## Discussion

Although earlier studies aimed to understand the infectiousness of asymptomatic carriers (8, 10, 11), the virological and immunological dynamics in these patients remain illusive. In this study, the clinical and laboratory features of asymptomatic, pre-symptomatic, and symptomatic SARS-CoV-2 infected patients were quantitatively described and analyze. To our knowledge, this was the first study to assess these characteristics for asymptomatic carriers.

As highlighted in recent studies, COVID-19-specific mortality is age-related, with deaths mainly occurring in patients over 60 years old; while young patients usually present with moderate or mild manifestations of the disease (12). The same was reported in this study, as the median age of asymptomatic patients was 30, and half of them were children who showed no or mild laboratory changes, including inflammation factors CRP and IL-6. One possible reason for asymptomatic COVID-19 cases to be more common in young adults is that a child or young person has a number of naíve immune cells, which can be easily educated by new antigens. By contrast, older populations have limited number of naíve immune cells, thus making them more susceptible to severe COVID-19 disease (13). Since that young people are more likely to be asymptomatic, thus they must be targeted in preventative efforts to assure proper precautions are taken and reduce the potential for transmission, especially in the context of schools.

Notably, there were 13 asymptomatic patients with abnormal chest CT scans, which was common in other studies as well (14, 15). This highlights the possibility of lung damage even if an individual is asymptomatic and not experiencing any other symptoms. Others have suggested that a chest CT be the first choice in the screening of close contacts and patients (16), which may be helpful for the early diagnosis and early treatment of infected patients. Especially as not all patients experience the signs and symptoms of COVID-19, using additional tools such as CT screenings may be more efficient and practical at capturing these high-risk transmitters.

For the viral dynamics of SARS-CoV-2, there were no obvious differences in the calculated initial Ct values among the three groups. Interestingly, as the fitted curve showed, the model-based initial viral load in nasopharyngeal swabs of asymptomatic carriers was the lowest, followed by symptomatic and pre-symptomatic patients. This may be related to the severity of disease, with symptomatic patients progressing to later stages of disease (17, 18). In addition, the RNA negative-conversion of asymptomatic carriers occurred 15-20 days after admission, while the RNA negative-conversion for pre-symptomatic and symptomatic patients occurred later. Thus, it could be concluded that asymptomatic carriers show an earlier viral clearance. Additionally noteworthy was that the viral RNA from two asymptomatic children was still detectable 50 days after admission, indicating continual shedding of the SARS-COV-2 virus. We also observed a re-appearance of SARS-CoV-2 virus RNA in eight asymptomatic patients after discharge, of whom had a positive RNA test up to 54 days after discharge, indicating they may continue shedding SARS-CoV-2 virus for a long time. Based on these observations, asymptomatic carriers showcased a large variation in viral dynamics. Therefore, vigilant control measures must be continued at this stage of the COVID-19 epidemic in order to avoid a resurgence caused by asymptomatic cases.

Asymptomatic, pre-symptomatic, and symptomatic patients all showed a rapid increase in IgG within seven days of symptom onset, confirming previously reported findings (19, 20). Importantly, most asymptomatic patients had constantly low levels of IgM, but high levels of IgG. As mentioned above, asymptomatic carriers show an earlier viral clearance. Thus, we guess that high and persistent IgM played a negative role in SARS-CoV-2 infection. Another reason may be asymptomatic patients did not experience an acute phase, or they may have already experienced the acute phase before they were discovered by active surveillance. Furthermore, half of the asymptomatic carriers are child and the vast majority of children had low levels of IgM. Future studies should investigate the intricate relationship of IgM, age, and clinical outcomes in order to improve control strategies.

This study has several limitations. First, this study was a single-center retrospective study, and multiple-center clinical observations are needed to evaluate the clinical, viral, and immunological characteristics of asymptomatic carriers with SARS-CoV-2 infection. Second, possible selection bias of enrolled patients may be present. As The Third People’s Hospital of Shenzhen is the only government mandated facility for the treatment of COVID-19, the study is generally representative of those infected with the disease. Third, it is unclear whether low levels of IgM play a protective role in asymptomatic carriers. Additional insights are needed which could strengthen the existing control measures in China.

In this study, asymptomatic carriers were found to be younger, with a lower initial viral load, early viral clearance,mild laboratory changes, mild chest CT manifestations, undetectable IgM and moderate levels of IgG. This sheds light on the management and potential immune mechanisms of viral clearance in asymptomatic carriers, useful tools that are needed to improve and strengthen existing care guidelines.

## Data Availability

The data used to support the findings of this study are included within the article

## Acknowledgements

We acknowledge the work and contribution of all the staffs from Shenzhen Third People’s Hospital. We thank all the patients involved in this study. We sincerely thank the biological sample bank of the Shenzhen Third People’s Hospital, clinical laboratory of SARS-CoV-2 detection of Institute of Hepatology and Beijing Wantai Biological Pharmacy Company for their helpful assistance.

## Reference

1. WHO. Coronavirus disease (COVID-19) outbreak situation. April 21, 2020. https://www.who.int/emergencies/diseases/novel-coronavirus-2019.

2. Mizumoto K, Kagaya K, Zarebski A, Chowell G. Estimating the asymptomatic proportion of coronavirus disease 2019 (COVID-19) cases on board the Diamond Princess cruise ship, Yokohama, Japan, 2020. Euro Surveill. 2020;25(10).

3. Hu Z, Song C, Xu C, Jin G, Chen Y, Xu X, et al. Clinical characteristics of 24 asymptomatic infections with COVID-19 screened among close contacts in Nanjing, China. Sci China Life Sci. 2020.

4. The epidemiological characteristics of an outbreak of 2019 novel coronavirus diseases (COVID-19) in China. Zhonghua liu xing bing xue za zhi = Zhonghua liuxingbingxue zazhi. 2020;41(2):145–51.

5. Mizumoto K, Chowell G. Transmission potential of the novel coronavirus (COVID-19) onboard the diamond Princess Cruises Ship, 2020. Infect Dis Model. 2020;5:264–70.

6. National Health Commission of the People’s Republic of China. Guideline of the prevention and control for novel coronavirus pneumonia (the sixth edition). March 7, 2020; http://www.nhc.gov.cn/iki/s3577/202003/4856d5b0458141fa9f376853224d41d7.shtml.

7. Bai Y, Yao L, Wei T, Tian F, Jin DY, Chen L, et al. Presumed Asymptomatic Carrier Transmission of COVID-19. JAMA. 2020.

8. Qian G, Yang N, Ma AHY, Wang L, Li G, Chen X, et al. A COVID-19 Transmission within a family cluster by presymptomatic infectors in China. Clin Infect Dis. 2020.

9. Tong ZD, Tang A, Li KF, Li P, Wang HL, Yi JP, et al. Potential Presymptomatic Transmission of SARS-CoV-2, Zhejiang Province, China, 2020. Emerg Infect Dis. 2020;26(5).

10. Wang Y, Liu Y, Liu L, Wang X, Luo N, Ling L. Clinical outcome of 55 asymptomatic cases at the time of hospital admission infected with SARS-Coronavirus-2 in Shenzhen, China. The Journal of infectious diseases. 2020.

11. Rothe C, Schunk M, Sothmann P, Bretzel G, Froeschl G, Wallrauch C, et al. Transmission of 2019-nCoV Infection from an Asymptomatic Contact in Germany. N Engl J Med. 2020;382(10):970–1.

12. Onder G, Rezza G, Brusaferro S. Case-Fatality Rate and Characteristics of Patients Dying in Relation to COVID-19 in Italy. Jama. 2020.

13. Weiskopf D, Weinberger B, Grubeck-Loebenstein B. The aging of the immune system. Transplant international: official journal of the European Society for Organ Transplantation. 2009;22(11):1041–50.

14. Meng H, Xiong R, He R, Lin W, Hao B, Zhang L, et al. CT imaging and clinical course of asymptomatic cases with COVID-19 pneumonia at admission in Wuhan, China. The Journal of infection. 2020.

15. Asadollahi-Amin A, Hasibi M, Ghadimi F, Rezaei H, SeyedAlinaghi S. Lung Involvement Found on Chest CT Scan in a Pre-Symptomatic Person with SARS-CoV-2 Infection: A Case Report. Tropical medicine and infectious disease. 2020;5(2).

16. Lin C, Ding Y, Xie B, Sun Z, Li X, Chen Z, et al. Asymptomatic novel coronavirus pneumonia patient outside Wuhan: The value of CT images in the course of the disease. Clinical imaging. 2020;63:7–9.

17. He X, Lau EHY, Wu P, Deng X, Wang J, Hao X, et al. Temporal dynamics in viral shedding and transmissibility of COVID-19. Nature medicine. 2020.

18. Wolfel R, Corman VM, Guggemos W, Seilmaier M, Zange S, Muller MA, et al. Virological assessment of hospitalized patients with COVID-2019. Nature. 2020.

19. Liu W, Liu L, Kou G, Zheng Y, Ding Y, Ni W, et al. Evaluation of Nucleocapsid and Spike Protein-based ELISAs for detecting antibodies against SARS-CoV-2. Journal of clinical microbiology. 2020.

20. Zhao J, Yuan Q, Wang H, Liu W, Liao X, Su Y, et al. Antibody responses to SARS-CoV-2 in patients of novel coronavirus disease 2019. Clinical infectious diseases: an official publication of the Infectious Diseases Society of America. 2020.

